# Using machine learning to design a short test from a full-length test of functional health literacy in adults - the development of a short form of the Danish TOFHLA

**DOI:** 10.1101/2023.01.05.23284233

**Authors:** Lisa Korsbakke Emtekær Hæsum, Simon Lebech Cichosz, Ole Kristian Hejlesen

## Abstract

**Introduction:** Patients are compelled to become more involved in shared decision making with healthcare professionals in the self-management of chronic disease and general adherence to treatment. Therefore, it is valuable to be able to identify patients with low functional health literacy so they can be given special instructions about the management of chronic disease and medications. However, time spent by both patients and clinicians is a concern when introducing a screening instrument in the clinical setting, which raises the need for short instruments for assessing health literacy that can be used by patients without the involvement of healthcare personnel. This paper describes the development of a short version of the full-length Danish TOFHLA (DS-TOFHLA) that is easily applicable in the clinical context and where the use does not require a trained interviewer.

**Materials and Methods:** Data were collected as a part of a large-scale telehomecare project (TeleCare North), which was a randomized controlled trial that included 1225 patients with chronic obstructive pulmonary disease.

The DS-TOFHLA was developed solely using an algorithm-based selection of variables and multivariable linear regression models. A multivariable regression model was developed using an exhaustive search strategy.

**Results:** The exhaustive search showed that the number of items in the full-length TOFHLA could be reduced from 17 numeracy items and 50 reading comprehension items to 20 reading comprehension items while maintaining a correlation of r = 0.90. A generic model-based approach was developed, which is suitable for development of short versions of the TOFHLA in other languages, including the original American version.

**Conclusions:** This study demonstrated how a generic model-based approach could be applied in the development of a short version of the TOFHLA, thereby reducing the 67 items to 20 items in the short version. Furthermore, this study showed that the inclusion of numeracy items was not necessary. The development of the DS-TOFHLA presents an opportunity to reliably identify patients with inadequate functional health literacy in approximately 5 minutes without involvement of healthcare personnel. The approach may be used in the development of short versions of any scaling questionnaire.

## 1. Introduction

Healthcare systems all over the world are developing in a way that compels patients to become more active in the management of their own health and disease – a development that changes the role of modern patients and the skills needed to navigate the healthcare system. Demographic changes resulting in more elderly people have led to increases in the burden of chronic diseases and put pressure on increasingly scarce healthcare resources (1). One strategy for overcoming this burden is to reduce the utilization of healthcare resources in the secondary sector by reducing the length of stay and placing more health care services in the primary sector, thus allowing more rehabilitation actions, where the goal is to have patients take control of their own life situation and health. This development focuses on both the ability to be an active part in shared decision making with healthcare professionals during the self-management of chronic diseases and general adherence to treatment, thus requiring that patients increase their understanding and application of health information (2) (3) (4). The World Health Organization (WHO), however, describes the presence of a paradox: increasing demands on the individual patient without the information and support necessary for making health-promoting choices (5). In the wake of this development in healthcare systems, the concept of health literacy (HL) is receiving increased attention. HL is defined by WHO as ‘the cognitive and social skills which determine the motivation and ability of individuals to gain access to, understand and use information in ways which promote and maintain good health’ (6).

As HL is a very complex concept, there is no universally accepted definition. A review by Sorensen et al. (7) found 17 different definitions of HL and 12 conceptual models. Even if there is no universally accepted definition, there are elements common to most definitions: obtaining, understanding, and applying health-related information. Don Nutbeam has described these three elements as functional (accessing health-related information), interactive (the ability to understand health-related information) and critical HL (the ability to actively use health-related information) (8). The myriad of definitions and conceptual models causes a lack of a universally agreed screening instrument to assess HL.

A recent review (9) aimed to identify the most optimal screening instrument for assessing HL in a clinical setting. The review identifies the S-TOFHLA (assessing basic numeracy and literacy skills related to healthcare) as the most widely used in the literature (used in nearly half of the studies), the REALM (medical word pronunciation test) as the second most used and NVS (the ability to identify and interpret basic text and mathematical calculations) as the third most used (9). It should be noted that the above-mentioned screening instruments are criticised for not capturing the complexity of HL (10,11) and as a result screening instruments that seek to capture the higher levels of HL has been developed (12,13). However, these instruments have a subjective approach focusing on self-experienced and self-rated abilities to perform tasks relevant to the management of health information. Thus, these more subjective screening instruments reflect self-evaluated skills in relation to the HL demands of specific hypothetical health-related situations (12,13). Further, the reliability of the subjective screening instruments can be questioned, as participants are prone to overrate their abilities, as a low level of HL is associated with shame and embarrassment (14). Despite the ongoing discussion about the nature of HL and how it should be assessed, the S-TOFHLA, REALM and NVS remain the most widely used in existing literature (9). The rapid technological development has added a new dimension to HL: the ability to consult electronic sources for information about health and use this information in relation to treatment and disease – referred to as e-health literacy (15). E-health literacy, and the assessment of this, is outside the scope of this paper.

A full-length Danish version of the full-length American TOFHLA (D-TOFHLA; D denoting Danish) was developed according to acknowledged guidelines with assessment of face validity, content validity, internal consistency etc. and has proven accurate in assessing functional health literacy (FHL) (16–18). Like the full-length American TOFHLA, the D-TOFHLA consists of two parts with a total of 67 items; the first part comprises 17 items assessing numeracy skills (e.g., prescription bottles, appointment cards that is administered by an interviewer) and the second part comprises 50 items assessing reading comprehension skills. As the D-TOFHLA (and the American) require involvement of an interviewer (e.g., healthcare personnel), it is, unfortunately, not suited for clinical routine use and it has thus primarily been used for research purposes (17,19).

This paper aimed to use machine learning to develop a short version of the D-TOFHLA (DS-TOFHLA; DS denoting Danish Short) that is easily applicable in the Danish clinical context and that does not require the involvement of an interviewer (healthcare personnel). This paper seeks to describe a generic model-based approach applied in the development of the short version of the D-TOFHLA that can also be used in the development of short versions of the TOFHLA in other languages or, in general, to develop short versions of full-length screening instruments.

## 2. Materials and Methods

The development of the DS-TOFHLA was based on the D-TOFHLA (17); the D-TOFHLA was created based on the original full-length American TOFHLA using the technique described by Beaton et al. (20). Similar to the original full-length American TOFHLA, the total score of the D-TOFHLA is divided into three levels: inadequate (0-59), marginal (60-74), and adequate (75-100); inadequate and marginal scores are regarded as ‘low FHL’ (19).

Motivated by a need to reduce the administration time and create an easier-to-use screening instrument, a short version of the original full-length American TOFHLA was designed: the S-TOFHLA (21). Due to the significant variations between the original American TOFHLA and the Danish version, it is not possible to translate and adapt the S-TOFHLA into a Danish version. The development of the S-TOFHLA was based on more subjective decisions and less on objective algorithm-based decisions, including linear regression models (LRM). The development of the DS-TOFHLA was based solely on an algorithm-based selection of variables and linear multiple regression (LMR). The classical method to obtain an unbiased evaluation, when building and testing models, is to have separate training and testing datasets, which can be accomplished by splitting a given dataset into a training set and a test set. In smaller datasets K-fold cross-validation is often used, which makes it possible to use almost the whole dataset for both training and testing while still avoiding bias. The present study used a special form of K-fold cross-validation, the leave-one-out cross-validation, where K is set to the number of samples (K = N), which, on the expense of more computation time during development, ensures the best possible use of the dataset (i.e., 100% of the dataset is used as training data and 100% as test data) (22).

The quality goal in the development of the DS-TOFHLA was expressed by two conditions. First, Pearson’s correlation coefficient between the DS-TOFHLA score (i.e., the predicted Danish TOFHLA total score) and the D-TOFHLA score should be at least 0.9 (r ≥ 0.9 being indicative of a very strong correlation). Second, if possible, the model should not contain numeracy items to eliminate the involvement of an interviewer.

### 2.1 Ethical approval

The trial has been presented to the Regional Ethical Committee for Medical Research in the North Denmark. The committee determined that no ethical approval was necessary.

### 2.2 Data material

The selection of items for the DS-TOFHLA was based on data from a previous large study that used the D-TOFHLA (16,18). Data were collected as a part of a large-scale telehomecare project, TeleCare North COPD (Chronic Obstructive Pulmonary Disease) (23). The 158 patients included in the study were relatively good representatives of Danish patients with chronic disease; the patients, in addition to COPD, had various chronic diseases: for example, 10% diabetes, 32% coronary heart disease, 5% mental health problem, 27% musculoskeletal disorder, and 5% cancer (24). Inclusion and exclusion criteria can be found in Table 1.

**Table 1.**
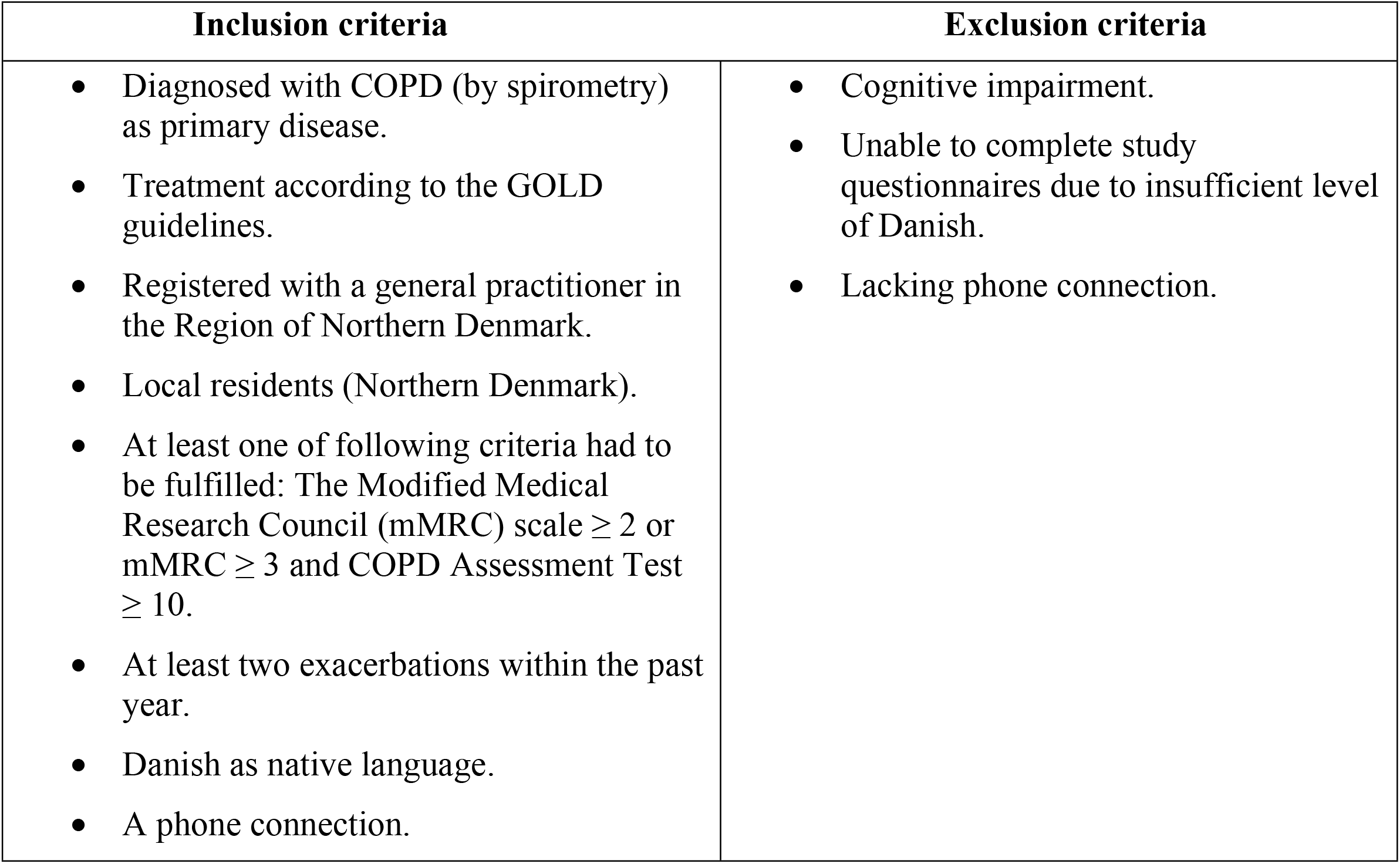
Inclusion and exclusion criteria

### 2.3 Development of prediction model

The D-TOFHLA is created as whole sentences, where one or more words are missing, and the participant is asked to select word/words that best complete a sentence. Therefore, before modelling the DS-TOFHLA, the 50 reading comprehension items in the D-TOFHLA were grouped into meaningful sets of items (items that create a whole sentence) to ensure that the intended meaning were maintained. This resulted in 19 sets: 5 sets with 1 item, 7 sets with 2 items, 1 set with 3 items, 3 sets with 4 items, 2 sets with 5 items, and 1 set with 6 items. The DS-TOFHLA was based on the following LMR equation:

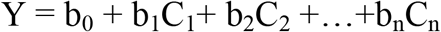

where Y is the total D-TOFHLA score, C_1_, C_2_,…, C_n_ are the reading comprehension items included in the DS-TOFHLA (from the D-TOFHLA), and b_0_,b_1,…,_ b_n_ are the regression coefficients that are adjusted to fit the model.

The model was developed using an exhaustive search strategy: for every model size (from a 1-item model and up until the quality criteria were met), all possible combinations of sets of comprehension items were tested. The root mean square error (RMS error or RMSE) between the DS-TOFHLA LRM and the D-TOFHLA score was used as the model fit criterion. After minimizing the RMSE, the model with the highest Pearson’s correlation coefficient was identified.

### 2.4 Validation of internal consistency

The internal consistency of the DS-TOFHLA was determined by using Cronbach’s alpha coefficient. An instrument is considered reliable if the Cronbach’s alpha exceeds a value of 0.7 (25). Item to scale correlations for all items were analysed using Pearson’s point-biserial correlation coefficient, where values of 0-0.2 are considered weak correlations, 0.2-0.5 are considered medium correlations, and 0.5-1 are considered high correlations (26).

### 2.5 The scoring system for DS-TOFHLA

Because of the DS-TOFHLA being based on a LRM that was used to predict the D-TOFHLA, the scoring system for the DS-TOFHLA was assumed to be similar to that of the D-TOFHLA: inadequate level: 0-59 points, marginal level: 60-74 points, adequate level: 75-100 points (17).

To assess the predicted scores of the DS-TOFHLA, a confusion matrix was used to illustrate the relation between the FHLs in the DS-TOFHLA and those in the D-TOFHLA.

The accuracy for predicting the three FHLs was calculated. In addition, the ability of the DS-TOFHLA to correctly detect ‘low FHL’ was assessed in accordance with the procedure described by Parker et al. (19).

### 2.6 Comparison with the short version of the original full-length American TOFHLA

The development of the S-TOFHLA was based on subjective decisions combined with some linear regression modelling, the latter being subjectively adapted to facilitate easy scoring (9,21). The S-TOFHLA includes the first 36 reading comprehension items and 4 numeracy items (item number 1, 4, 5, and 8) from the original full-length TOFHLA. The reading comprehension items were weighted by assigning a score of 2 points to each and the numeracy items were weighted by assigning a score of 7 points to each. Hence, the maximum score for the 36 reading comprehension items and the 4 numeracy items was 72 and 28, respectively, yielding a maximum total score of 100, which is the same as for the full-length American TOFHLA (19).

For comparison, a subset of the items in the D-TOFHLA was selected in accordance with the subset used in the S-TOFHLA (the first 36 reading comprehension items and numeracy items 1,4,5, and 8) and, using the same weighting as in the S-TOFHLA (2 and 7 respectively), a Danish mirror version of the S-TOFHLA was constructed. Thus, the ‘D-36-4-TOFHLA’ was based on 40 items from the D-TOFHLA combined in the following LMR equation (reading comprehension, R, and numeracy, N):

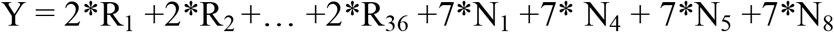

Pearson’s correlation coefficient between the D-36-4-TOFHLA score and the D-TOFHLA score, for the 158 COPD patients recruited from the TeleCare North cohort, was calculated.

Most studies using the S-TOFHLA, have chosen to omit the numeracy items (9). Even though this prose only version simplifies the test, it may also introduce additional bias. A second Danish mirror version of the ‘Prose S-TOFHLA’, omitting the 4 numeracy items, was constructed. Thus, the ‘D-36-0-TOFHLA’ was based on the following simplified LMR equation:

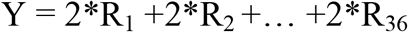

Using the same 158 COPD patients, Pearson’s correlation coefficient between the D-36-0-TOFHLA score and the D-TOFHLA score was calculated.

## 3. Results

The basic demographic characteristics of the 158 participants can be found in Table 2. The mean age was 69.6 years (SD: 9.53). The basic characteristics were relatively balanced, except for educational level; 20% of the participants completed high school or higher education, and 80% completed elementary school or skilled work.

**Table 2.** Basic demographics of the participants.

|  |
| --- |
| **Age, mean (SD):** 69.6 (9.53) |
| **Sex, n (%)** |
| - Male: 76 (48) |
| - Female: 82 (52) |
| **Civil status, n (%)** |
| - Living alone: 76 (48) |
| - Married or living with a partner: 82 (52) |
| **Educational level, n (%)** |
| - Elementary school: 53 (34) |
| - High school: 11 (7) |
| - Higher education: 20 (13) |
| - Skilled: 74 (47) |
| **Health literacy level, n (%)** |
| - Inadequate: 39 (25) |
| - Marginal: 33 (21) |
| - Adequate: 86 (54) |
| **Health literacy score, mean (SD, range):** 71.6 (18.7, 13-100) |

The exhaustive search showed that the number of items in the D-TOFHLA could be reduced to 20 reading comprehension items and that there was no need for numeracy items. The sets of reading comprehension items were item 2-3, item 13-14, item 18-21, item 23-25, item 37-41, and item 42-45, each set corresponding to a sentence in the Danish TOFHLA leading to the following regression model:

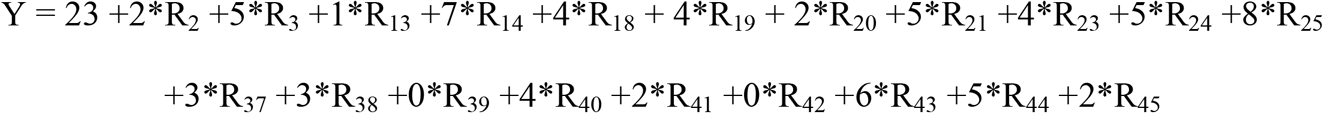

Two examples from the six sentences in the DS-TOFHLA are listed in *Appendix A*. The maximum time for administration could be reduced from 22 minutes (10 minutes for numeracy items and 12 minutes for comprehension items) to 5 minutes (12*20/50 minutes).

Figure 1 illustrates the development of the best possible model performance as a function of the number of reading comprehension items in the model. The figure shows that the best model with only one item had a correlation coefficient of 0.6, and the best model with 20 items had a correlation coefficient of 0.9. The scatter plot in Figure 2 illustrates the relation between the DS-TOFHLA and the D-TOFHLA for the best model with 20 items. The correlation coefficient was 0.90 (SD 0.01), and the correlation was highly significant (p<0.001).

**Figure 1.**
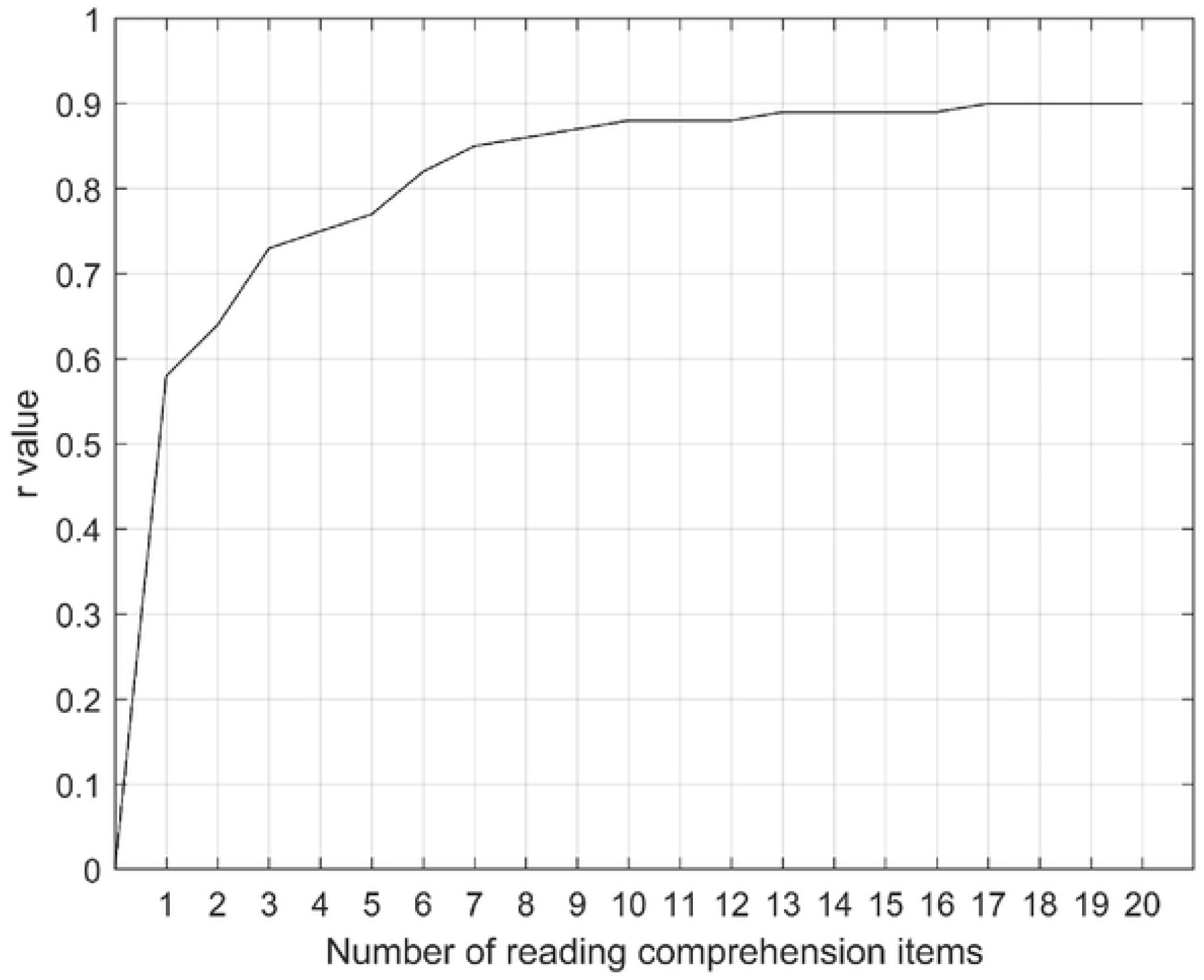
The development of the best possible model performance as a function of the number of reading comprehension items in the model.

**Figure 2.**
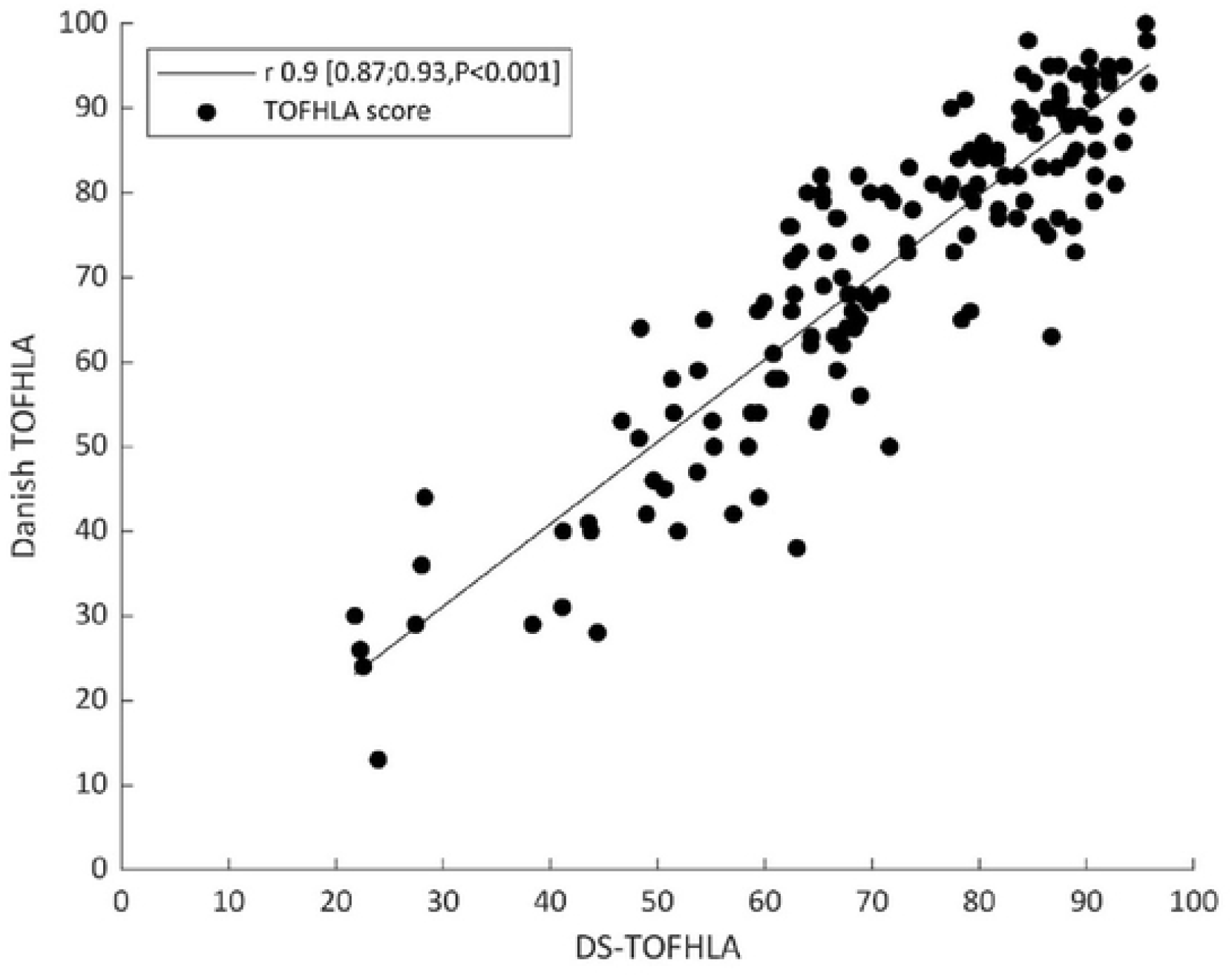
The relation between the DS-TOFHLA and the Danish TOFHLA for the best model with 20 items.

The internal consistency measured by Cronbach’s alpha was 0.885. This indicated that the reliability of the DS-TOFHLA was acceptable (>0.7 as set by Houser (25)). Item to scale correlations were assessed for all 20 items using Pearson’s point-biserial correlation coefficient; 12 items showed a high correlation and 8 items showed a medium correlation. The analysis of the Pearson’s point-biserial correlation coefficient showed significant positive correlations between all 20 items and the scale (p < 0.01).

Table 3 shows a confusion matrix illustrating the ability of the model to correctly predict each participant’s HL level. In the confusion matrix, for 126 out of 158 participants, the prediction was correct; for 32 out of the 158 participants, the prediction was off by one level; and no prediction was off by more than one level.

**Table 3.** Confusion matrix for prediction of health literacy levels

|  |  |  |  |
| --- | --- | --- | --- |
| True inadequate | 28 | 11 | 0 |
| True marginal | 2 | 26 | 5 |
| True adequate | 0 | 14 | 72 |
|  | Predicted inadequate | Predicted marginal | Predicted adequate |

The accuracy of the prediction of the inadequate level (lowest level) (i.e., inadequate vs. marginal or adequate) was 92%. The accuracy of the prediction of the marginal level (middle level) (i.e., marginal vs. adequate or inadequate) was 80%. The accuracy of the prediction of the adequate level (highest level) (i.e., adequate vs. marginal or inadequate) was 88%, which can also be expressed as the accuracy of the prediction of ‘low FHL’ as defined by Parker et al. (19).

To enable a comparison with the performance of the S-TOFHLA, the relation between the D-36-4-TOFHLA (the Danish mirror version of S-TOFHLA) score and the D-TOFHLA score, for the 158 COPD patients recruited from the TeleCare North cohort, is illustrated in Figure 3. Pearson’s correlation coefficient between the two scores was 0.90. Likewise, the relation between the D-36-0-TOFHLA (the Danish mirror version of the Prose S-TOFHLA) score and the D-TOFHLA score, for the 158 COPD patients recruited from the TeleCare North cohort, is illustrated in Figure 3B. Pearson’s correlation coefficient between the two scores was 0.85. Table 4 gives an overview of the various model versions of TOFHLA.

**Table 4.** Overview of the various model versions of TOFHLA listing language (English, Danish), number of reading comprehension items (prose items), number of numeracy items, indication of most relevant use (research, clinical), maximum time for administration (minutes), and, for the short Danish versions, Pearson’s correlation coefficient with D-TOFHLA (r).

| **Model version** | **Language** | **No. of read. comp. items** | **No. of num. items** | **Mostly research use** | **Suited for clinical use** | **Max. time for administration** | **Pearson's correlation coefficient with D-TOFHLA** |  |
| --- | --- | --- | --- | --- | --- | --- | --- | --- |
| TOFHLA | E | 50 | 17 | + |  | 22 |  | Original American version |
| S-TOFHLA | E | 36 | 4 |  | ? | 12 |  | Short American version |
| Prose S-TOFHLA | E | 36 |  |  | + | 9 |  | Simplified 'prose only' short American version |
| D-TOFHLA | D | 50 | 17 | + |  | 22 |  | Danish cross-cultural adaptation of TOFHLA |
| D-36-4-TOFHLA | D | 36 | 4 |  | ? | 12 | 0.90 | Danish mirror version of S-TOFHLA |
| D-36-0-TOFHLA | D | 36 |  |  | + | 9 | 0.85 | Danish mirror version of Prose S-TOFHLA |
| DS-TOFHLA | D | 20 |  |  | + | 5 | 0.90 | Danish short version developed using LMR |

**Figure 3.**
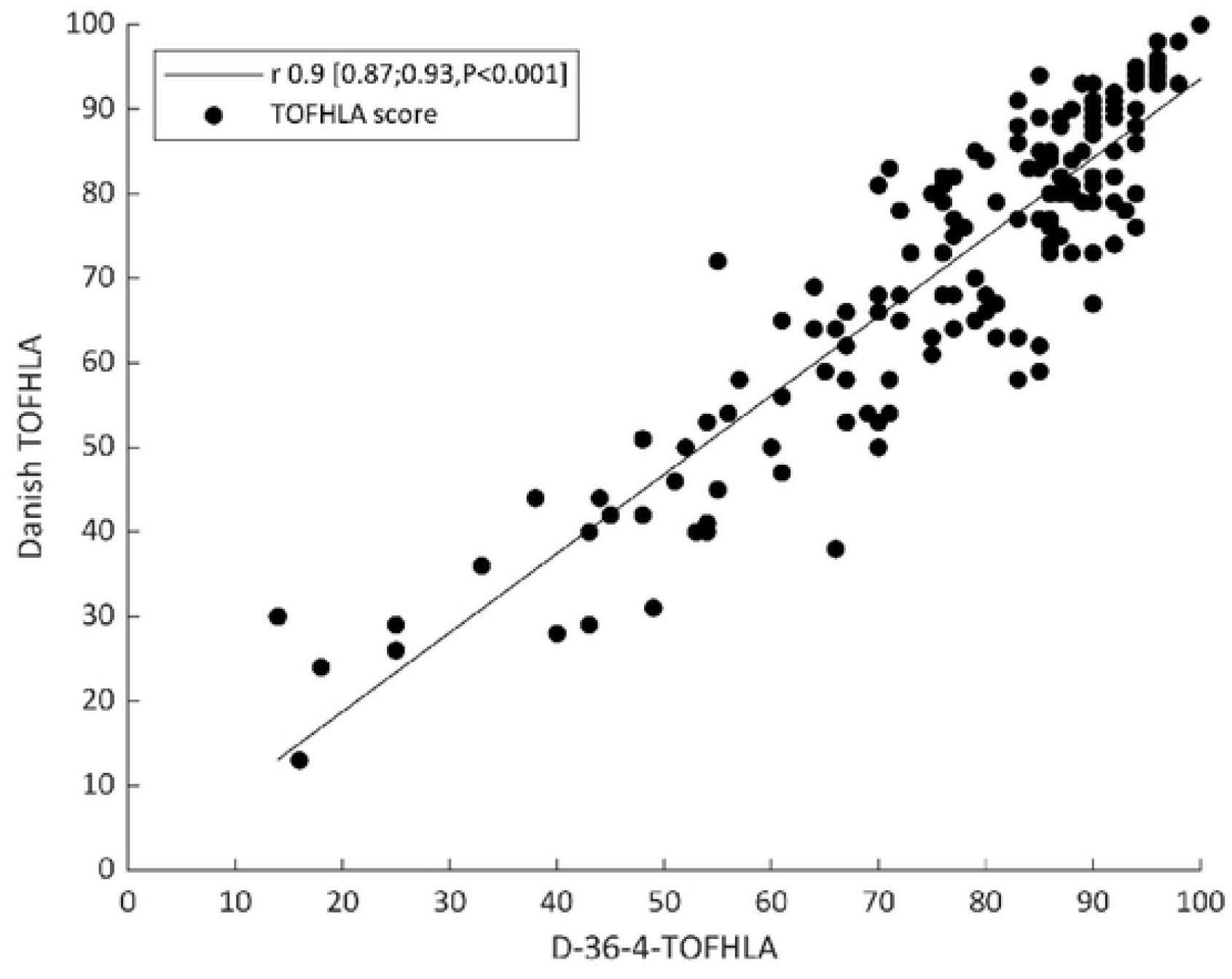
The relation between the Danish mirror version of the S-TOFHLA (D-36-4-TOFHLA) and the Danish TOFHLA.

**Figure 4.**
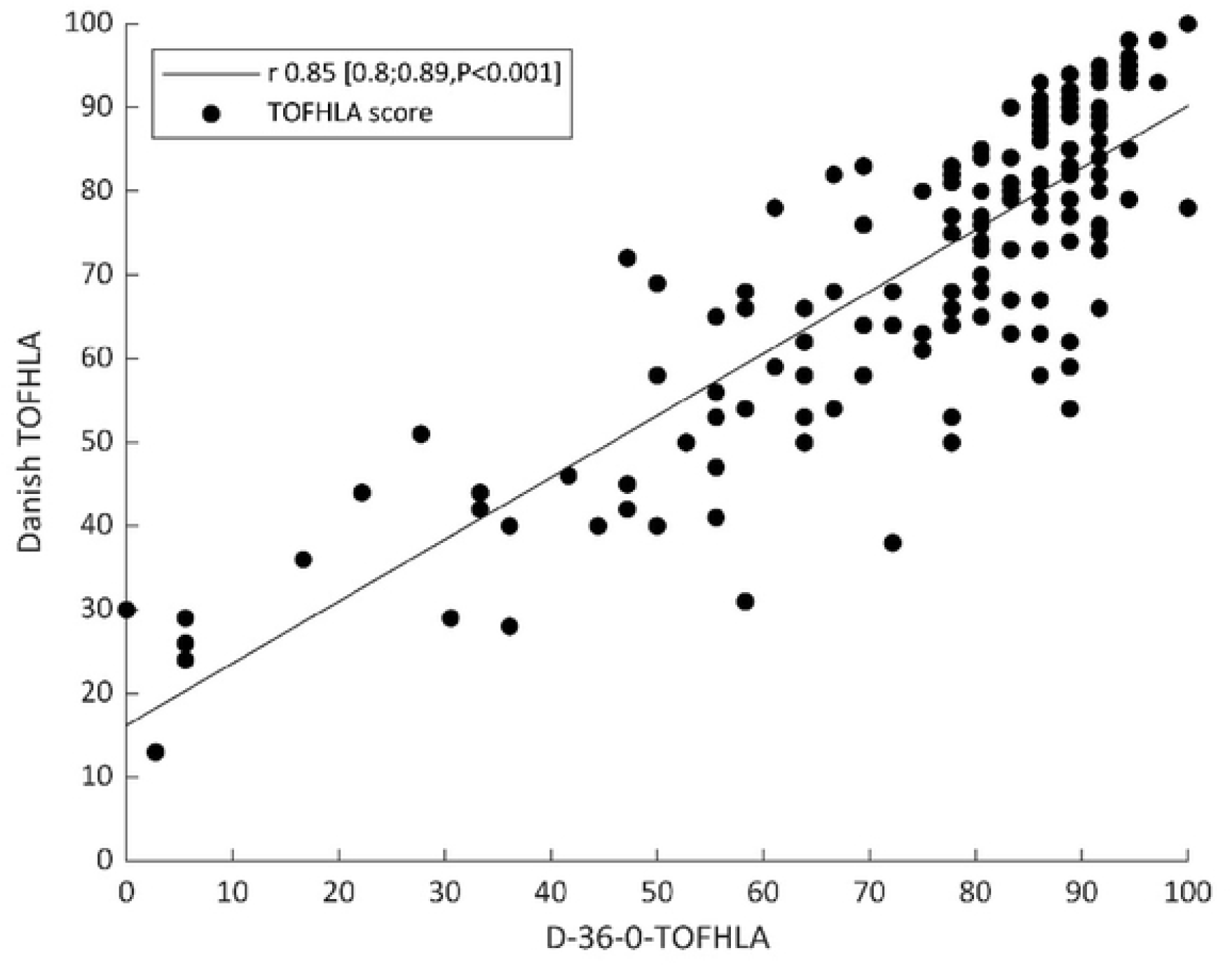
The relation between the Danish mirror version of the Prose S-TOFHLA (D-36-0-TOFHLA) and the Danish TOFHLA.

## 4. Discussion

The aim of this study was to investigate the use of machine learning to develop a short test of FHL in adults (the DS-TOFHLA) that can be used in the development of short versions of the TOFHLA in various languages, including the original version of the American TOFHLA in English. In addition to investigating the machine learning approach, this study also addressed the problem that, to the authors’ knowledge, there are no efficient, suitable, and objective screening instruments for assessing HL in a clinical setting in Denmark and other non-English speaking countries.

A review has shown that most studies using the S-TOFHLA chose to omit the numeracy items to simplify the test and make it usable in a clinical setting (9). In our study, statistical analyses showed that inclusion of numeracy items was not necessary to meet the chosen quality goal of the study. By including only 20 reading comprehension items, it was possible to create a short version of the D-TOFHLA where the use does not require a trained interviewer. Therefore, in contrast to only using this instrument for research purposes, the DS-TOFHLA is also applicable as a screening instrument for the clinical setting. In addition, the maximum time for administration was reduced from 22 minutes to 5 minutes.

For comparison, an assessment of the performance of the S-TOFHLA was performed using Danish mirror versions of the short American versions. Both the S-TOFHLA and the Prose S-TOFHLA (without numeracy items) was assessed, the latter being the most widely used (9). Pearson’s correlation coefficient, when comparing scores from both the Danish mirror version of S-TOFHLA and the DS-TOFHLA with scores from D-TOFHLA, was 0.90. This indicates that DS-TOFHLA, even though it, as opposed to S-TOFHLA, does not include numeracy items and therefore is easily applicable in a clinical setting, has the same level of performance as S-TOFHLA. Furthermore, the maximum time for administration of DS TOFHLA is only 5 minutes compared to 12 minutes for S-TOFHLA. Pearson’s correlation coefficient when comparing scores from the Danish mirror version of the Prose S-TOFHLA with scores from D-TOFHLA was 0.85. This indicates that the Prose S-TOFHLA, which has omitted numeracy items and therefore as opposed to the S-TOFHLA is more applicable in a clinical setting, is inferior in performance to DS-TOFHLA. Furthermore, it should be noted that the maximum time for administration of the Prose S-TOFHLA is significantly longer than for DS-TOFHLA (9 minutes and 5 minutes respectively).

The DS-TOFHLA was developed solely using an algorithm-based selection of variables and LRMs. A major strength of this method was that the design principles were founded on objective algorithm-based decisions, and the LRM used for development selected the items from the D-TOFHLA that led to the most accurate predictions of the level of FHL. The reading comprehension items in both the D-TOFHLA and the American TOFHLA are ordered by increasing difficulty in readability level and thus, it is reasonable to assume that the more difficult items are most accurate in predicting the functional level of HL. In this regard, it should be noted that only 4 of 20 items selected for the DS-TOFHLA by the algorithm herein were from the first and easiest part of the reading comprehension items. The 36 items assessing reading comprehension in the S-TOFHLA is primarily from the first and middle part (lowest difficulty) and none from the latter and most difficult part. In line with this, the regression model presents quite different weights to the selected items for the DS-TOFHLA, e.g., C42 = 0 and C43 = 6. In comparison, the development of the American S-TOFHLA seems based on a subjective decision to include the first 36 items, without explicitly considering if these items contribute to the most accurate prediction of the FHL or considering the ascending difficulty in readability.

The accuracy of the prediction of the three levels was ranged from 80%-92%; the middle level had the lowest accuracy, which can be explained by the fact that this level is defined by a relatively narrow range of 60-74 points. The prediction of the lowest and highest levels was a one-sided classification, whereas the prediction of the middle layer was two-sided. It should be noted that the prediction of ‘low FHL’ (i.e., inadequate or marginal vs. adequate scores), as defined by Parker et al (19), had an accuracy of 88%.

The DS-TOFHLA was based on a predictive LRM and should not be regarded as a de novo questionnaire, and it, therefore, should not go through the same rigorous evaluation. Instead, the focus should be on developing a short questionnaire with the best possible predictions of the validated full-length questionnaire. Likewise, it makes sense to develop the prediction model in the DS-TOFHLA based on the same data that was used to develop and validate the full-length Danish TOFHLA (16,18). However, further work might be carried out to test the model on other datasets and other types of patients. Alternatives to LRMs might be considered. However, even though other classification methods such as neural networks or various clustering methods might have yielded higher correlation coefficients with 20 reading comprehension items, the results from using such models would be harder to explain both to experts in the field and to clinicians using the HL score. The study used an exhaustive search strategy. Many studies have applied forward or backward selection strategies. Even though such strategies reduce the computational burden significantly, they also lead to the risk of not finding the best model and were therefore not used in the study.

## 5. Conclusion

This study demonstrated how a generic model-based approach could be applied in the development of a short version of the TOFHLA, thereby reducing the 67 items in the full-length version to 20 items. Furthermore, this study showed that the inclusion of numeracy items was not necessary to meet the chosen quality goal of a Pearson’s correlation coefficient ≥0.9, resulting in a short version of TOFHLA where the use does not require a trained interviewer. The work was based on Danish data and a validated Danish full-length version of TOFHLA. The generic model-based approach used herein may also be used in the development of short versions of the TOFHLA in other languages and in the development of short versions of any scaling questionnaire.

## Data Availability

The data underlying the results presented in the study are available from Aalborg University

## References

1. The World Health Organization. Noncommunicable diseases [Internet]. 2022. Available from: https://www.who.int/news-room/fact-sheets/detail/noncommunicable-diseases

2. Jordan JE, Osborne RH. Chronic disease self-management education programs: challenges ahead. Medical Journal of Australia [Internet]. 2007 Jan 15;186(2):84–7. Available from: https://onlinelibrary.wiley.com/doi/abs/10.5694/j.1326-5377.2007.tb00807.x

3. Coleman K, Austin BT, Brach C, Wagner EH. Evidence on the Chronic Care Model in the new millennium. Health Aff. 2009 Jan;28(1):75–85.

4. Joosten EAG, DeFuentes-Merillas L, de Weert GH, Sensky T, van der Staak CPF, de Jong CAJ. Systematic review of the effects of shared decision-making on patient satisfaction, treatment adherence and health status. Psychother Psychosom. 2008 May;77(4):219–26.

5. Kickbusch I, Pelikan JM, Apfel F, Tsouros AD, World Health Organization. Regional Office for Europe. Health literacy: the solid facts. 73 p.

6. Nutbeam D. Health promotion glossary. Health Promot. 998;1(1):113–27.

7. Sørensen K, van den Broucke S, Fullam J, Doyle G, Pelikan J, Slonska Z, et al. Health literacy and public health: A systematic review and integration of definitions and models. Vol. 12, BMC Public Health. 2012.

8. Nutbeam D. Health literacy as a public health goal: a challenge for contemporary health education and communication strategies into the 21st century. Health Promot Int [Internet]. 2000;15(3):259–67. Available from: http://heapro.oxfordjournals.org.ezproxy.library.wisc.edu/content/15/3/259

9. Duell P, Wright D, Renzaho AMN, Bhattacharya D. Optimal health literacy measurement for the clinical setting: A systematic review. Vol. 98, Patient Education and Counseling. Elsevier Ireland Ltd; 2015. p. 1295–307.

10. Nielsen-Bohlman L, Panzer AM, Kindig D a. Health Literacy: A Prescripton to End Confusion. Intstitute of Medicine. 2004.

11. Baker DW. The meaning and the measure of health literacy. J Gen Intern Med. 2006;21(8):878–83.

12. Sørensen K, van den Broucke S, Pelikan JM, Fullam J, Doyle G, Slonska Z, et al. Measuring health literacy in populations: illuminating the design and development process of the European Health Literacy Survey Questionnaire (HLS-EU-Q). BMC Public Health [Internet]. 2013;13:948. Available from: http://www.pubmedcentral.nih.gov/articlerender.fcgi?artid=4016258&tool=pmcentrez&rendertype=abstract

13. Osborne RH, Batterham RW, Elsworth GR, Hawkins M, Buchbinder R. The grounded psychometric development and initial validation of the Health Literacy Questionnaire (HLQ). BMC Public Health. 2013;13(1).

14. Haun J, McCormack L, Valerio M, Sorensen K. Health Literacy Measurement: Health Literacy Measurement: An inventory and descriptive summary of 52 instruments. J Health Commun. 2014;0730(September 2015).

15. Norman CD, Skinner HA. eHealth Literacy: Essential Skills for Consumer Health in a Networked World. J Med Internet Res. 2006;8.

16. Hæsum LKE, Ehlers LH, Hejlesen OK. The long-term effects of using telehomecare technology on functional health literacy: results from a randomized trial. Public Health. 2017 Sep 1;150:43–50.

17. Emtekær Hæsum LK, Ehlers L, Hejlesen OK. Validation of the Test of Functional Health Literacy in Adults in a Danish population. Scand J Caring Sci. 2015;29(3).

18. Korsbakke Emtekaer Haesum L, Ehlers L, Hejlesen OK. Interaction between functional health literacy and telehomecare: Short-term effects from a randomized trial. Nurs Health Sci. 2016 Sep 1;18(3):328–33.

19. Parker RM, Baker DW, Williams M v, Nurss JR. The test of functional health literacy in adults: a new instrument for measuring patients’ literacy skills. J Gen Intern Med. 1995;10(10):537–41.

20. Beaton DE, Bombardier C, ¶#§, Guillemin F, Ferraz MB. Guidelines for the Process of Cross-Cultural Adaptation of Self-Report Measures. Vol. 25, SPINE.

21. Baker DW, Williams M v, Parker RM, Gazmararian J a, Nurss J. Development of a brief test to measure functional health literacy. Patient Educ Couns. 1999;38(1):33–42.

22. Refaeilzadeh P, Tang L, Liu H. Encyclopedia of Database Systems. Cross-Validation. Liu L, Özsu MT, editors. New York, NY: Springer New York; 2016.

23. Udsen FW, Lilholt PH, Hejlesen O, Ehlers LH. Effectiveness and cost-effectiveness of telehealthcare for chronic obstructive pulmonary disease: study protocol for a cluster randomized controlled trial. Trials [Internet]. 2014;15(1):178. Available from: http://www.pubmedcentral.nih.gov/articlerender.fcgi?artid=4039321&tool=pmcentrez&rendertype=abstract

24. Udsen FW, Lilholt PH, Hejlesen O, Ehlers L. Cost-effectiveness of telehealthcare to patients with chronic obstructive pulmonary disease: Results from the Danish TeleCare North’ cluster-randomised trial. BMJ Open. 2017 May 1;7(5).

25. Houser J. Nursing research Reading, Using and Creating Evidence. 2nd ed. Boston: Jones & Barlett Publishers; 2011.

26. Everitt B. The Cambridge Dictionary of Statistics. 2nd ed. Cambridge: Cambridge University Press; 2002.

